# The occurrence and persistence of Imposter Phenomenon in clinicians and medical students in Wales

**DOI:** 10.1101/2025.06.09.25329299

**Authors:** Jasmine Pattarukuzhyil Jose, Adam Barter-Jones, Umakant Dave, Menna Brown, Alice E. Hoon

## Abstract

**Purpose of the article:** Imposter phenomenon (IP) is characterised by persistent feelings of inadequacy and self-doubt. IP among medical students and clinicians has been linked to poor mental health effects, with implications for patient care.

**Materials and methods:** The present study surveyed incidence of IP in medical students and clinicians in Wales. An online 81-item survey was developed, consisting of validated inventories that measured IP, burnout, perfectionism, and wellbeing, and bespoke questions. Quantitative data was analysed using independent t-tests, one-way ANOVA, and Pearsons correlation coefficient; qualitative data was analysed thematically.

**Results:** Two hundred and fifteen participants completed the survey. Approximately 75% of participants reported frequent to intense levels of IP. Mean IP scores did not differ significantly between medical students and clinicians, or between career stage. Women reported higher levels of IP than men. IP was positively correlated with perfectionism and burnout, and negatively correlated with wellbeing, which increased with career level. Qualitative data revealed participant difficulties with comparisons to others and doubts about competence.

**Conclusions:** IP is highly prevalent among medical students and clinicians in Wales, and does not alleviate with clinical experience. These results highlight the importance of understanding the complex relationship between IP, burnout, perfectionism, and wellbeing.

## Introduction

Imposter phenomenon (IP) encompasses feelings of intellectual fraudulence or doubt regarding your own success and achievement, and individuals may feel fearful of being exposed as a fraud (Clance & Imes, 1978). IP is particularly prevalent in competitive professions with high achieving peers or in high pressure settings, such as medicine (Cohen & McConnell, 2019; Huecker et al., 2024). Within medical professionals, IP can manifest as feeling inadequate or being overly critical of oneself, therefore feeling the need to maintain a façade of professionalism with a detriment to learning from clinical experiences (Chodoff, Conyers, Wright, & Levine, 2023). High prevalence of IP among medical students and clinicians has been linked to negative mental health (Thomas & Bigatti, 2020) and it has been suggested that the self-doubt and insecurity associated with IP may have implications for patient care, clinical decision-making, and overall professional wellbeing (Clark et al., 2022).

Existing literature suggests that IP may also be associated with increased levels of burnout and perfectionism. Shanafelt et al. (2022) reported that physicians had higher levels of IP than the general working population, and that high levels of IP were associated with increased burnout and suicidal ideation. Participants with IP also reported decreased job satisfaction (Shanafelt et al., 2022). Issues associated with IP may already present for some individuals when in medical school. Houseknecht et al. (2019) reported that IP increased in students during their time at medical school, whilst wellbeing decreased. Campos et al. (2022) found a positive correlation between IP severity with depression and burnout in medical students. Futhermore, students with a previous history of depression and anxiety were more likely to experience IP (Campos, et al., 2022).

Perfectionism may also be associated with IP. Perfectionism can serve both adaptive or maladaptive functions, and is a common trait found in medical professionals (Thomas & Bigatti, 2020). Individuals with high levels of perfectionism may have high personal expectations, however this can manifest as perceiving vulnerability or help seeking to be a weakness (Shanafelt et al., 2022). It may also be exacerbated by a fear of causing a medical error or negative patient outcomes (Robertson & Long, 2018). A recent review article (Thomas & Bigatti, 2020) reported significant correlations between IP, perfectionism, as well as poor mental health outcomes such as depression, anxiety, suicidal ideation, burnout.

To our knowledge, there is a paucity of research as to whether IP changes during a medical career. Clark et al. (2021) reported that there was no association with age or years of work with IP amongst mental health professionals. Chodoff et al (2023) reported transition periods, to roles with increased responsibility and expectation, led to increased anxiety on self-adequacy. LaDonna et al. (2018) noted that self-doubt was reported in even experienced clinicians, however, this research was qualitative in nature and IP was not measured.

Villwock et al (2016) reported a significant association of female gender with IP compared to males. Thomas and Bigatti (2020) suggests environments with less female role models may exacerbate IP in females, alongside any inner turmoil experienced working away from traditional gender norms. Implicit gender-based bias and disparities faced by female clinicians may negatively impact confidence and feelings of IP (Cawcutt et al., 2021).

Despite growing interest in IP, there is a paucity of research conducted in the UK. Hence, this study aimed to assess the associated factors and effects of IP on medical students and clinicians based in Wales (UK) with the aim to develop targeted interventions.

## Materials and methods

### Ethics

This study was approved by Swansea University Medical School Research Ethics Committee (Ref: 1 2023 7882 6910).

### Study design

The present study used a survey design. Microsoft Forms was used to host the 81-item online survey. The questionnaire consisted of the following five sections: preliminary and demographic data, imposter phenomenon, perfectionism, burnout, wellbeing and additional information. Each section included an open free-text question where participants could add additional comments.

### Outcome measures

The Clance Imposter Phenomenon Scale (CIPS) assessed the presence of IP (Clance, 1985). This 20-item scale uses a 5-point Likert scale with total scores ranging from 20 to 100. While there is no universally established cutoff score, values exceeding 60 and 80 are indicative of moderate and severe imposter tendencies, respectively (Paladugu, Wasser, & Donato, 2021).

The Frost Multidimensional Perfectionism Scale-Brief (FMPS-Brief; Burgess et al., 2016) is an 8-item using 5-point Likert scales measuring perfectionism through two sub-sections: Striving (S) and Evaluative Concerns (EC) with four items each (Simon, 2020).

The Oldenburg Burnout Inventory (OBI) is a 16-item scale assessing burnout using a 5-point scale. The OBI analyses two dimensions of burnout: disengagement and exhaustion, with scores >2.1 and >2.25 respectively, indicating high levels of burnout (Bellanti, et al., 2021).

The Warwick-Edinburgh Mental Wellbeing Scale (WEMWBS) consists of 14-items to measure mental wellbeing using 5-point Likert scales (Maheswaranet al., 2012). Item scores are summed to produce a total score between 14 and 70, with higher total score implying higher level of mental wellbeing. A score of 43.5 or below is considered a screening threshold for depression.

### Setting and participants

Medical students studying in Welsh medical schools and clinicians working in Wales deanery were invited to participate in the survey between October 2023 and February 2024. The survey was distributed via email, through postgraduate centres and institution announcements. Participation was voluntary.

### Procedure

The recruitment email included a link to the survey. On clicking the link, participants were first presented with the Participant Information Sheet and only those who confirmed they had read this, met the eligibility criteria, and provided informed consent could proceed to the survey. Consent was recorded through tick boxes hosted on the survey platform. Upon consenting, the survey then commenced. At the end of the survey, participants were provided with a debrief which included signposting to mental health support.

### Data analysis

Survey data was accessed on Saturday 11^th^ February 2024. All data was anonymous, however some participants voluntarily provided an e-mail address if they were willing to be invited to take part in a later interview. This data was stored separately to their survey data. Data was analysed Microsoft Excel and IBM SPSS. Descriptive statistics summarised the demographic data. Independent sample t-tests and one-way ANOVAs compared mean CIPS scores by demographic variables (e.g. gender, stage of career). Pearson correlation coefficients assessed the relationships between IP, burnout, perfectionism, and wellbeing. Free text responses were coded and analysed thematically. This process was informed by the six stages of reflective thematic analysis outlined by Braun and Clarke (2006).

## Results

### Participants

Two hundred and fifteen participants completed the survey, including 127 clinicians and 87 medical students (see Table 1). Clinicians were split into three sub-groups based on stage of career: Junior (Foundation Year between FY1-FY3), Intermediate (Specialist Training, Core Training, Core Surgical Training, GP Speciality Training, Internal Medical Training, and Specialist Registrar), and Senior (Speciality and Associate Specialist and consultants).

**Table 1.** Participant demographics.

| Demographic | Frequency (%) |
| --- | --- |
| Ethnicity |  |
| Arab | 6 (2.8) |
| Asian | 22 (10.2) |
| Black | 5 (2.3) |
| Mixed | 11 (5.1) |
| White | 170 (79) |
| Prefer not to say | 1 (0.5) |
| **Gender** |  |
| Female | 136 (63.3) |
| Male | 72 (33.5) |
| Other | 4 (1.9) |
| Prefer not to say | 3 (1.4) |
| **Stage of career** |  |
| Medical student | 87 (40.5) |
| Junior clinician | 59 (27.6) |
| Intermediate clinician | 52 (24.2) |
| Senior clinician | 16 (7.5) |

Overall, 75% of participants reported experiencing frequent to intense levels of IP, with only 5% of the sample experiencing the lowest level. Amongst clinicians, IP prevalence was highest for the intermediate career group, followed by junior level, then senior level (See Table 2).

**Table 2.** Percentage of Imposter Phenomenon by demographic.

| Level of IP | Overall (%) | Medical student | Junior | Intermediate | Senior | Male | Female |
| --- | --- | --- | --- | --- | --- | --- | --- |
| **Few** | 5.1 | 2.3 | 1.7 | 9.6 | 18.8 | 9.7 | 2.9 |
| **Moderate** | 20 | 23 | 23.7 | 9.6 | 25 | 22.2 | 17.6 |
| **Frequent** | 46.5 | 43.7 | 49.2 | 50 | 43.8 | 44.4 | 47.8 |
| **Intense** | 28.4 | 31 | 25.4 | 30.8 | 12.5 | 23.6 | 31.6 |

### Statistical analyses

#### Correlation between IP with perfectionism, burnout, and wellbeing

Pearsons correlations were conducted between IP and perfectionism, burnout, and wellbeing for all participants (see Table 3). A strong positive correlation was found between IP and perfectionism, a weak positive correlation was found between IP and burnout, and a moderately negative correlation was observed between IP and wellbeing (Figure 1).

**Table 3.**
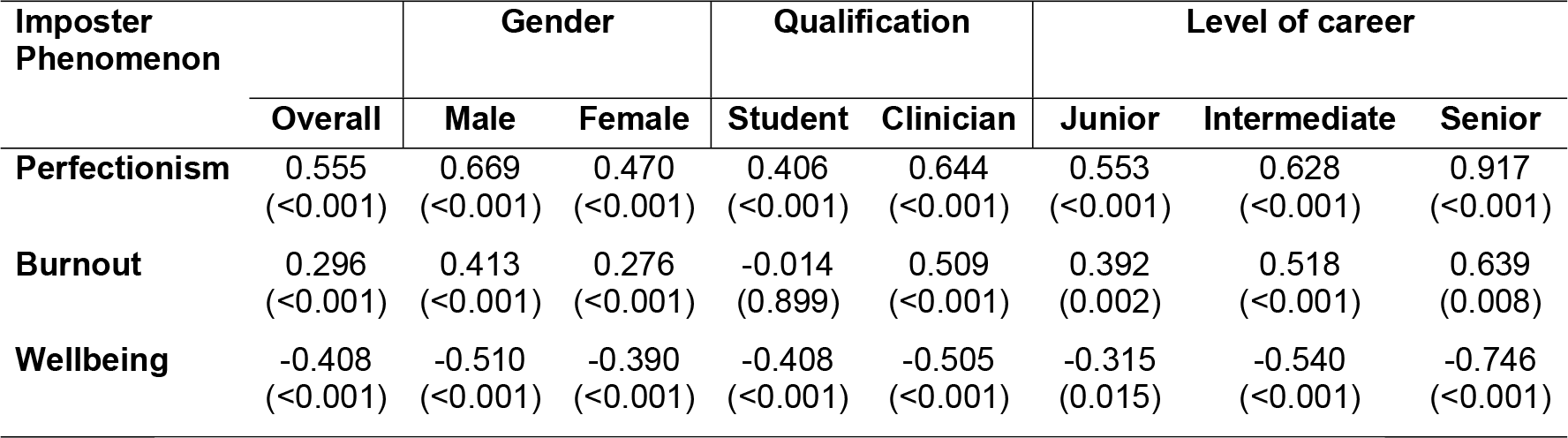
Correlation coefficient of imposter phenomenon with perfectionism, burnout and wellbeing by demographic (p value in brackets).

**Figure 1.**
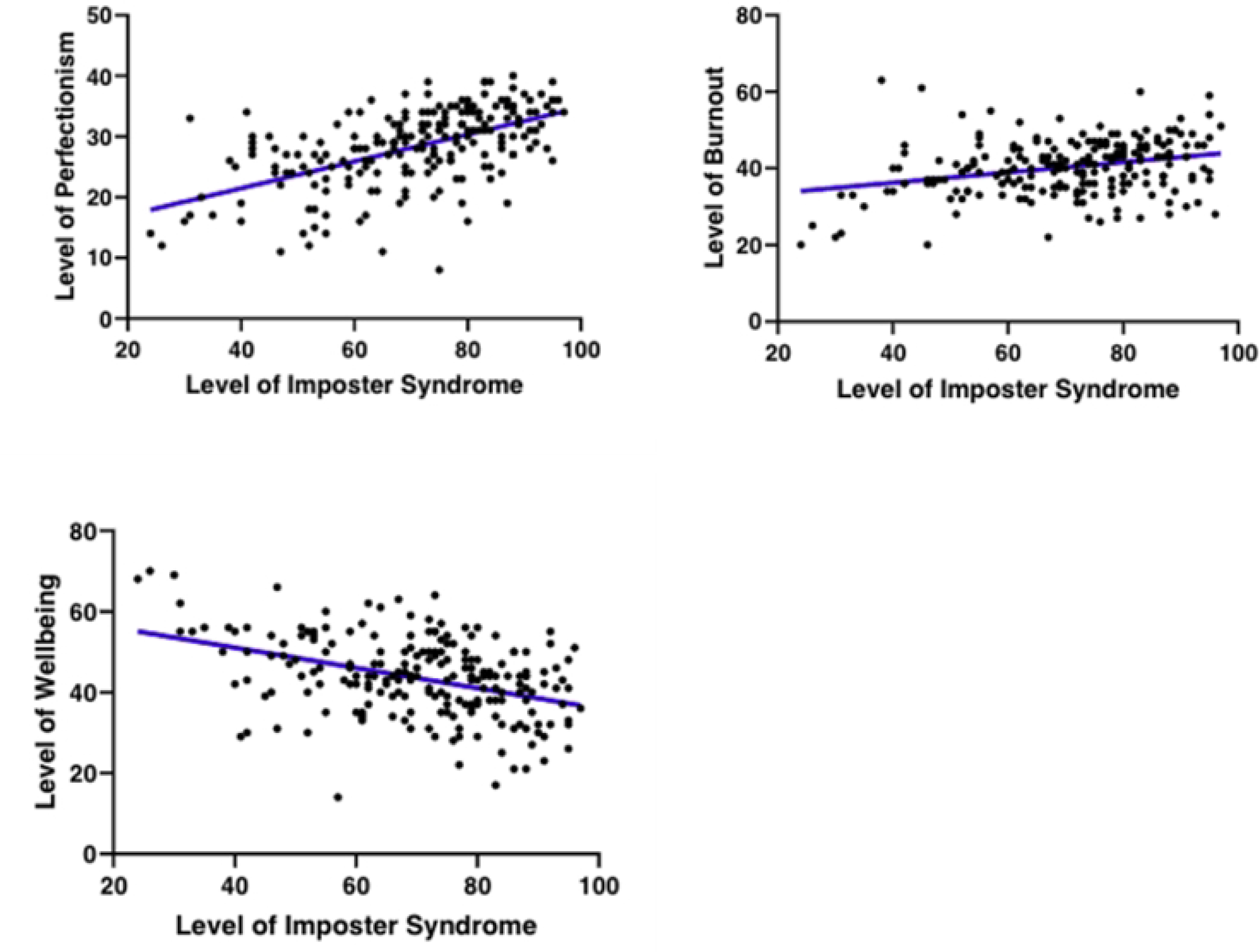
Correlation of Imposter Phenomenon with Perfectionism, Burnout, and Wellbeing for all participants.

#### Analysis between clinicians and medical students

Further analyses were conducted to analyse data by demographic. In clinicians, the correlations between IP with perfectionism, burnout and wellbeing were stronger than medical students. For clinicians, there was a strong positive correlation between IP with both perfectionism (Figure 2) and burnout, and a strong negative correlation with wellbeing.

**Figure 2.**
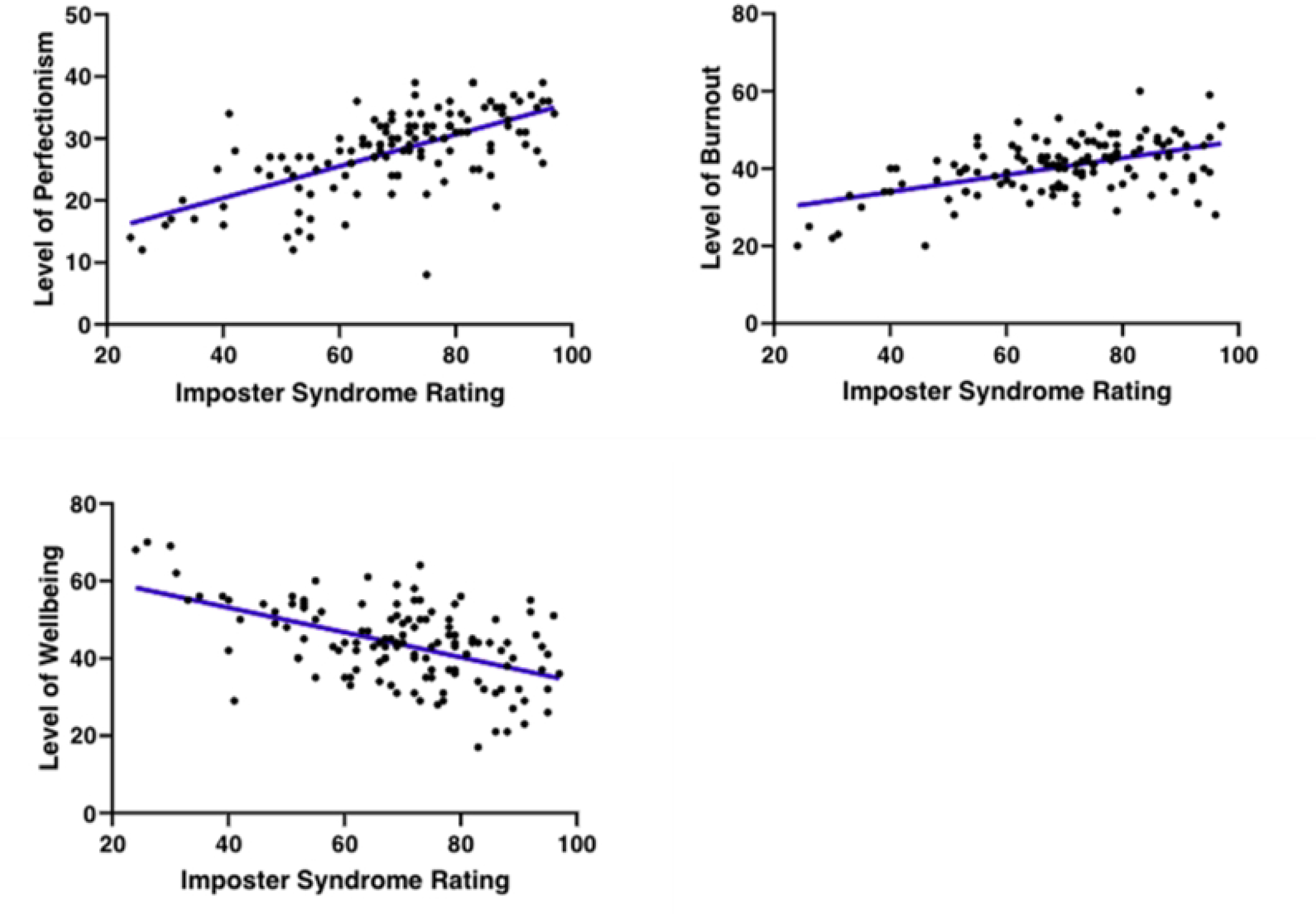
Correlation of imposter phenomenon with perfectionism, burnout and wellbeing for clinicians.

For medical students IP demonstrated a moderately positive correlation with perfectionism (Figure 3) and a weak negative correlation with wellbeing. There was no correlation for burnout.

**Figure 3.**
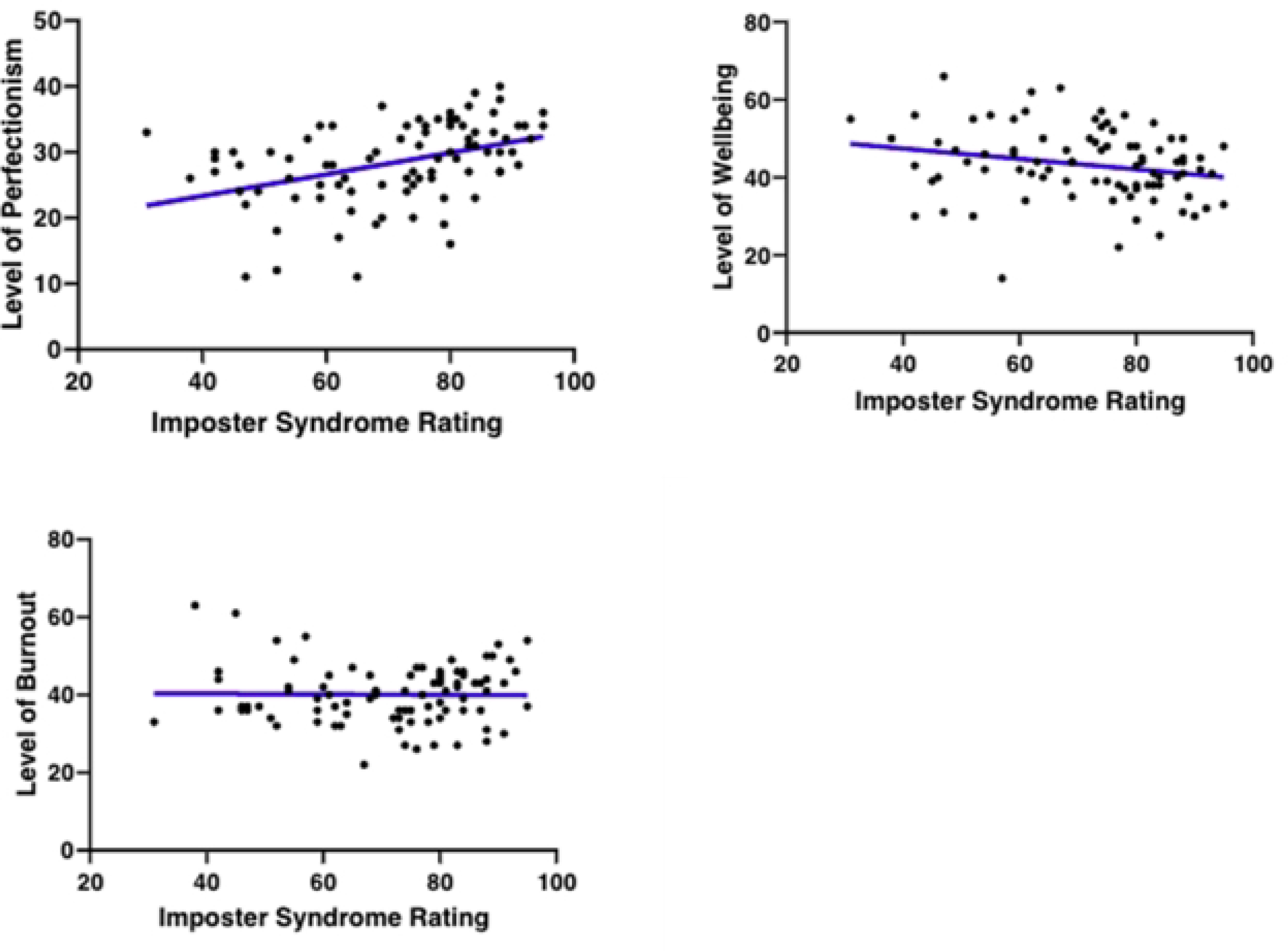
Correlation of imposter phenomenon with perfectionism, burnout and wellbeing in medical students.

#### Analysis by stage of career

One participant who did not specify their stage of career was omitted from this analysis. When data was analysed depending on the stage of career, a relatively strong positive correlation between IP and perfectionism was observed in junior clinicians, a moderate positive correlation between IP and burnout, and a moderate negative correlation between IP and wellbeing (see Table 3). Among intermediate clinicians, the correlation between IP and perfectionism was stronger than junior clinicians. Strong correlations were found between IP and burnout and wellbeing in intermediate clinicians. For senior clinicians, the correlation between IP and perfectionism was near perfect (r=0.917). The correlation between IP, burnout and wellbeing was stronger among for senior clinicians, suggesting that the relationship strengthens with increasing seniority among clinicians. Although no assumptions are required to measure a linear relationship using Pearson’s correlation, the small sample of senior participants means there is a lack of certainty that the results are representative of the wider population (Schober et al., 2018).

Four one-way ANOVAs were conducted between IP, burnout, wellbeing, and perfectionism across different stages of career. There was no statistically significant difference for stage of career on IP [F(2, 124) = 2.38, p = 0.97] or perfectionism [F(2, 124) = .388, p = 0.714]. However, there were significant differences in burnout and wellbeing between senior (M=35.3, SD=8.78) and junior (M=42.4, SD=6.23) clinician groups [F(2, 124) = 7.75 p<0.001], which indicated higher levels of burnout among junior compared to senior clinicians. Similarly, with wellbeing, a significant difference was found between the senior (M=48.9, SD=8.98) and junior (M=41.7, SD=8.49) groups [F(2, 124) = 3.54 p=0.03], suggesting lower levels of wellbeing among junior clinicians compared to senior clinicians. The mean IP score for clinicians was slightly higher than for medical students, however the difference was not statistically significant. Independent t-tests revealed that no statistically significant differences for perfectionism, burnout or wellbeing between clinicians and medical students (see Table 4).

**Table 4.** Independent T-test between clinicians and medical students.

|  | Clinician |  | Medical students |  | t | p |
| --- | --- | --- | --- | --- | --- | --- |
|  | M | SD | M | SD |  |  |
| **Imposter Phenomenon (CIPS)** | 69.7 | 16.4 | 71.1 | 15.43 | 0.631 | 0.264 |
| **Perfectionism** | 28.01 | 6.54 | 28.44 | 6.21 | 0.482 | 0.315 |
| **Burnout (OBI)** | 40.5 | 7.03 | 40.1 | 7.61 | 0.373 | 0.355 |
| **Wellbeing (WEMWBS)** | 43.6 | 10.27 | 43.29 | 9.22 | 0.229 | 0.409 |

#### Analysis by gender

Among male participants the correlations were stronger than for females. Specifically, IP demonstrated a strong positive correlation with perfectionism and a negative correlation for wellbeing, indicating that male participants reporting higher levels of IP had higher levels of perfectionistic tendencies and lower levels of wellbeing. IP exhibited a moderately positive correlation with burnout. A moderate positive correlation was observed between IP and perfectionism for females, and there was a weak positive correlation between IP and burnout. There was a moderate negative correlation with wellbeing.

An independent sample t-test revealed that males reported significantly lower levels of IP, perfectionism, and burnout than females (Table 5). Males reported higher levels of wellbeing, however, these differences were not statistically significant.

**Table 5.** Independent t-tests by gender.

|  | Males |  | Females |  | t | p |
| --- | --- | --- | --- | --- | --- | --- |
|  | M | SD | M | SD |  |  |
| **Imposter Phenomenon (CIPS)** | 67.4 | 17.78 | 72.12 | 14.85 | -2.044 | 0.021 |
| **Perfectionism** | 27.0 | 6.98 | 28.8 | 5.94 | -1.879 | 0.031 |
| **Burnout (OBI)** | 37.9 | 7.044 | 41.2 | 6.78 | -3.313 | 0.001 |
| **Wellbeing (WEMWBS)** | 45.4 | 10.56 | 42.9 | 8.94 | 1.584 | 0.057 |

### Qualitative data

Fifty-nine free text comments were recorded in the survey. Responses provided context to the survey data and highlighted the real-world context of IP for clinicians and medial students alike. In the IP and perfectionism sections of the survey, many participants wrote about their tendency to internalise criticism and make unfavourable comparisons of themselves to others. Participants reported feeling that their success was associated with chance or luck rather than personal competence, which contributed to feelings of inadequacy, and self-doubt. They noted feeling anxious about making mistakes and maintaining high standards that they perceived were expected of them, which sometimes culminated in pressure and a fear of failure. Lastly, they mentioned pressure to maintain a reputation for excellence, both personally and within their family, leading to self-imposed high expectations and a fear of disappointment. For example;

> *“I have extremely high standards of myself but not the persistence or aptitude to achieve those goals”*
>
> *“I often feel like I will never know enough to be safe or competent no matter how much I learn”*

In the burnout section of the survey, participants described feeling emotionally exhausted, experiencing energy depletion, fatigue, and low motivation which impacted their ability to enjoy life outside of work. Participants also reported a variety of factors which exacerbated feelings of burnout, such as lack of support from colleagues, job rotations, workload, and the deteriorating condition of health services.

> *“These feelings have worsened since I qualified 8 years ago as the health service and the care we can provide has deteriorated. Similarly, since Covid these feelings have dramatically worsened*.”

Similar comments were reported for wellbeing, where current events or personal circumstances sometimes influenced their overall mood and outlook on life. For example:

> *“I find that current affairs really affect my world view at any particular time. I get frustrated with individual patients and staff members with minor problems when people in Israel and Gaza are suffering unimaginable pain with limited resources*.*”*

Taken together, the qualitative and quantitative data indicate that perfectionism, burnout, wellbeing, and imposter phenomenon are related.

## Discussion

The present findings indicate that rates of imposter phenomenon are high in both clinicians and medical students in Wales, with 75% of participants reporting frequent to intense IP levels. Furthermore, there were significant positive correlations between IP with perfectionism and burnout, and a significant negative correlation between IP and wellbeing. There were no differences in IP between clinicians at different stages of their career, indicating that it is not something that eases with clinical experience.

Previous studies on IP have reported lower prevalence rates than were found in the present study, for example Alsaleem, et al. (2021) reported a rate of 41.2% in medical students. However, this study used the Young Imposter Scale (Villwock et al., 2016), which has dichotomous Yes or No categories for IP, whereas CIPS categorises IP into low, moderate, frequent, and intense. This makes it difficult to draw true comparisons. A USA-based study by Shanafelt et al. (2022) which used CIPS, also reported much lower prevalence of IP with only 23% of clinicians reporting frequent to intense IP.

Whilst it may be assumed that IP would decrease with career progression, the present study did not support this as there was no significant difference in IP between junior and senior clinicians. Furthermore, a stronger correlation between IP with perfectionism and wellbeing was found for consultants compared to those in junior roles. This continuation of IP may in part be due to the persistent nature of difficult thoughts which can be lifelong and resistant to change. Additionally, as a doctor progresses through their medical career, responsibility increases which may bring new challenges and therefore feelings of self-doubt do not diminish. The demanding nature of the profession combined with the pressure to excel, and a fear of mistakes, may all contribute to a persistence of IP.

In the present study, burnout had almost no correlation with IP in medical students which is contrary to what has previously been reported (see Villwock et al., 2016; Qureshi et al., 2017). A moderate correlation, however, was seen for clinicians which increased with stage of training. Addressing burnout should be considered a significant priority due to its links to adverse effects on mental health, professional development, and risks to patient care (Dyrbye & Shanafelt, 2015). The increasing correlation across different levels of career underscore the importance of interventions targeted at specific career stages.

Perfectionism had the strongest correlation to IP, both in medical students and clinicians at all levels of career. The qualitative data suggested that setting unattainable standards and experiencing anxiety, frustration or disappointment when those standards are not met, may contribute IP. This cycle of striving for perfection whilst fearing failure makes for a problematic combination.

Existing literature on gender differences of IP have shown conflicting results with one meta-analysis suggesting that the IP is higher in women (Price, Holcomb, & Payne, 2024) whereas others have found more nuanced results (see Badawyet al., 2018). Our findings found that women experienced greater prevalence of IP than men. However, despite the higher prevalence of IP in women, the correlation between IP and with stress, burnout, and wellbeing were weaker than those in men. Whilst it is not possible to make any assertations about causation here, perhaps IP is an indication of poorer mental health more generally in men, whereas women can experience IP at higher levels but without the effects of stress and burnout. Therefore, interventions aimed at stress and burnout may not necessarily lead to improvement in IP in women. Further research exploring the relationship between gender and IP which looks to establish causal factors is needed.

### Study Limitations

Several limitations must be considered when reviewing the results of the current study. The sample size accounted for about 2% of Welsh medical students and hospital doctors therefore may not be considered representative. Furthermore, there was variation in distribution among different career levels with a relatively small number of senior clinicians in the sample which may limit the ability to generalise for this subgroup.

A further issue is that self-selection bias was possible due to voluntary participation, therefore perhaps only those clinicians and students that are aware of their feelings of IP were more inclined to participate. Other contextual factors may have influenced the participants respondents as at the time of survey distribution NHS Wales doctors had chosen to strike due to NHS conditions and pay.

Finally, the study’s confinement to Wales may be a limitation. Wales has a distinct cultural and institutional context due to healthcare being a devolved matter and also has population demographic differences and longer waiting lists compared to England (GSS Coherence Team, 2024).

## Conclusion

In conclusion, prevalence of IP is high in Welsh clinicians and medical students, and it remains consistent across all stages of career, suggesting it does not decrease with time, career or expertise; Equally it is more prevalent in females. Thus, whilst interventions targeting associated factors may benefit all individuals, understanding unique experiences of women and medical students may identify effective interventions to support these specific populations.

## Data Availability

All anonymised data will be made available on OSF.io

## Acknowledgements

The authors would like to acknowledge Dr Pauline Rose Clance for permission to use the Clance Imposter Phenomenon Scale (CIPS) in the survey. As per copyright permission, see note below:

From The Imposter Phenomenon: When success makes you feel like a fake (pp. 20-22), by P.R. Clance, 1985, Toronto: Bantam Books. Copyright 1985 by Pauline Rose Clance. Reprinted by permission. Do not reproduce without permission from Pauline Rose Clance.

## Author contributions *(conceived/design/data collection/data analysis/main manuscript writing and revision)*

JPJ – conception, design, data collection, data analysis, manuscript writing and revision

ABJ - data collection, data analysis, manuscript writing and revision

AH – design, data collection, data analysis, manuscript writing and revision MB - design, data analysis, manuscript writing and revision

UD – conception, design, manuscript revision

All authors have read and approved the submitted version and agree to be personally accountable for contributions and questions related to the accuracy or integrity of the work.

## Ethics declaration

The Swansea University Medical School Ethics Committee at Swansea University approved the study (1 2023 7882 6910) on 19/10/2023. All participants in the study provided written informed consent. There was no funding for this research.

